# 12-lead Electrocardiogram in Hospitalized COVID 19 Patients

**DOI:** 10.1101/2021.01.29.21250407

**Authors:** Mohamed Shokr, Omar Chehab, Mustafa Ajam, Manmohan Singh, Said Ashraf, John Dawdy, Mohit Pahuja, Vivek Reddy, Ahmed Subahi, M. Chadi Alraies, Luis Afonso, Randy Lieberman

**Affiliations:** Department of Cardiology, Wayne State University, Detroit, MI, USA; Department of Internal Medicine, Wayne State University, Detroit, MI, USA; Department of Pulmonary and Critical Care Medicine, Oregon Health and Science University, Portland, OR, USA

## Abstract

COVID-19 pandemic resulted in considerable morbidity and mortality. We analyzed 345 Electrocardiograms of 100 COVID-19 patients admitted to our tertiary care center in Detroit, during the initial month of Covid-19. Findings were correlated with mortality, cardiac injury and inflammatory markers. Our cohort included 61% males and 77% African Americans. The median age and BMI were 66 years (57-74) and 31 kg/m^2^ (26.1-39), respectively. We observed atrial arrhythmias in 29% of the patients (17% new onset), First degree heart block in 12%, ST-T segment changes in 17%, S1Q3T3 pattern in 19%, premature ventricular complexes in 23%, premature atrial complexes in 13%, Q waves in 27%, T wave inversion in 42% of the cases. While presence of premature atrial complexes or left atrial abnormality correlated with mortality (P = 0.02 & 0.03, respectively), other findings did not show significant correlation in this small cohort of patients.

## Manuscript

COVID-19 pandemic resulted in considerable morbidity and mortality (1). We analyzed 345 Electrocardiograms of 100 COVID-19 patients admitted to our tertiary care center in Detroit, during the initial month of Covid-19. Findings were correlated with mortality, cardiac injury and inflammatory markers (Tables 1, 2S &3S supplementary index).

**Table 1:** Correlation of Electrocardiogram (EKG) findings with mortality, intensive care unit (ICU) admission, Cardiac injury (High sensitivity(Hs) Troponin > 17 ng/L) and Median C-reactive protein (175 mg/L)

| EKG finding | Prevalence | Survival |  | P-value | ICU admission |  | P-value | Hs Troponin |  | P-value | Median CRP |  | P-value |
| --- | --- | --- | --- | --- | --- | --- | --- | --- | --- | --- | --- | --- | --- |
|  |  | yes | No |  | No | Yes |  | ≤17 | >17 |  | <175 | ≥175 |  |
| Transient 1<sup>st</sup> °HB | 6% | 1.9% | 10.4% | 0.2 | 10.8% | 3.2% | 0.055 | 9.5% | 6.4% | 0.7 | 9.1% | 2.3% | 0.2 |
| Persistent 1<sup>st</sup> °HB | 6% | 5.8% | 6.3% |  | 0.0% | 9.5% |  | 4.8% | 2.1% |  | 4.5% | 9.1% |  |
| T wave inversion | 42% | 46.2% | 37.5% | 0.2 | 43.2% | 41.3% | 0.5 | 61.9% | 36.2% | 0.04 | 36.4% | 52.3% | 0.09 |
| Premature ventricular complexes | 23% | 15.4% | 31.3% | 0.050 | 21.6% | 23.8% | 0.5 | 14.3% | 23.4% | 0.3 | 25.0% | 18.2% | 0.3 |
| Premature atrial | 13% | 5.8% | 20.8% | 0.02 | 8.1% | 15.9% | 0.2 | 4.8% | 12.8% | 0.3 | 15.9% | 9.1% | 0.3 |
| complexes |  |  |  |  |  |  |  |  |  |  |  |  |  |
| Rhythm on admission |  |  |  | 0.9 |  |  | 0.059 |  |  | 0.3 |  |  | 0.2 |
| Sinus rhythm | 56% | 57.7% | 54.2% |  | 70.3% | 47.6% |  | 66.7% | 46.8% |  | 61.4% | 45.5% |  |
| Sinus Tachycardia | 33% | 30.8% | 35.4% |  | 18.9% | 41.3% |  | 28.6% | 42.6% |  | 27.3% | 43.2% |  |
| Atrial fibrillation | 11% | 11.5% | 10.4% |  | 10.8% | 11.1% |  | 4.8% | 10.6% |  | 11.4% | 11.4% |  |
| Rhythm during admission |  |  |  |  |  |  |  |  |  |  |  |  |  |
| Sinus bradycardia | 13% | 17.3% | 8.3% | 0.1 | 24.3% | 6.3% | 0.01 | 9.5% | 10.6% | 0.6 | 18.2% | 9.1% | 0.1 |
| Sinus tachycardia | 81% | 78.8% | 83.3% | 0.4 | 67.6% | 88.9% | 0.01 | 90.5% | 80.9% | 0.3 | 72.7% | 90.9% | 0.02 |
| Left atrial abnormality | 5.1% | 9.8% | 0% | 0.03 | 5.6% | 4.8% | 0.6 | 0.0% | 4.3% | 0.5 | 2.4% | 9.1% | 0.1 |
| Q waves |  |  |  | 0.5 |  |  | 0.02 |  |  | 0.6 |  |  | 0.1 |
| Q waves (Inferior) | 22% | 21.2% | 22.9% |  | 21.6% | 22.2% |  | 23.8% | 25.5% |  | 18.2% | 29.5% |  |
| (Anterior) | 2% | 3.8% | 0% |  | 5.4% | 0.0% |  | 0.0% | 2.1% |  | 2.3% | 0.0% |  |
| ≥2 territories | 3% | 3.8% | 2.1% |  | 8.1% | 0.0% |  | 0.0% | 6.4% |  | 6.8% | 0.0% |  |
| Low voltage | 16% | 9.6% | 22.9% | 0.06 | 13.5% | 17.5% | 0.4 | 14.3% | 14.9% | 0.6 | 18.2% | 11.4% | 0.27 |
| LVH | 15% | 17.3% | 12.5% | 0.3 | 16.2% | 14.3% | 0.5 | 9.5% | 19.1% | 0.3 | 6.8% | 25.0% | 0.01 |
| Prolonged QTc | 37% | 36.5% | 37.5% | 0.5 | 29.7% | 41.3% | 0.2 | 42.9% | 34.0% | 0.3 | 31.8% | 45.5% | 0.1 |
| S1Q3T3 | 19% | 17.3% | 20.8% | 0.4 | 24.3% | 15.9% | 0.2 | 23.8% | 25.5% | 0.6 | 18.2% | 22.7% | 0.3 |

**Table 2S:** Correlation of EKG findings with Mortality, ICU admission, Cardiac injury and elevated Brain Natriuretic peptide (BNP) level (> 100 pg/ml)

| EKG finding | N=100 | Survivors |  | P-value | ICU admission |  | P-value | Cardiac injury |  | P-value | BNP |  | P-value |
| --- | --- | --- | --- | --- | --- | --- | --- | --- | --- | --- | --- | --- | --- |
|  |  | yes | No |  | No | Yes |  | No | Yes |  | ≤100 | >100 |  |
| 1<sup>st</sup> degree heart block (HB) | 12% | 7.7% | 16.7% | 0.1 | 10.8% | 12.7% | 0.5 | 14.3% | 8.5% | 0.37 | 11.8% | 12.5% | 0.7 |
| Transient 1<sup>st</sup> degree HB | 6% | 1.9% | 10.4% | 0.2 | 10.8% | 3.2% | 0.055 | 9.5% | 6.4% | 0.7 | 5.9% | 4.2% | 0.9 |
| Persistent 1<sup>st</sup> degree HB | 6% | 5.8% | 6.3% |  | 0.0% | 9.5% |  | 4.8% | 2.1% |  | 5.9% | 8.3% |  |
| Transient ST elevation | 5% | 3.8% | 6.3% | 0.4 | 2.7% | 6.3% | 0.5 | 4.8% | 6.4% | 0.4 | 5.9% | 4.25 | 0.7 |
| Persistent ST elevation | 3% | 1.9% | 4.2% |  | 0.0% | 4.8% |  | 4.8% | 2.1% |  | 5.9% | 0.0% |  |
| ST depression | 8% | 11.5% | 4.2% |  | 8.1% | 7.9% |  | 14.3% | 6.4% |  | 11.8% | 8.3% |  |
| Early repolarization | 1% | 1.9% | 0% |  | 2.7% | 0.0% |  | 4.8% | 0.0% |  | 0.0% | 4.2% |  |
| T wave inversion | 42% | 46.2% | 37.5% | 0.2 | 43.2% | 41.3% | 0.5 | 61.9% | 36.2% | 0.04 | 41.2% | 50.0% | 0.4 |
| T wave inversion (inferior) | 5% | 3.8% | 6.3% | 0.5 | 2.7% | 6.3% | 0.4 | 4.8% | 6.4% | 0.04 | 0.0% | 12.5% | 0.4 |
| (anterior) | 18% | 21.2% | 14.6% |  | 16.2% | 19.0% |  | 14.3% | 19.1% |  | 23.5% | 25.0% |  |
| (Lateral) | 2% | 3.8% | 0% |  | 5.4% | 0.0% |  | 4.8% | 2.1% |  | 5.9% | 0.0% |  |
| (≥ 2 territories) | 17% | 17.3% | 16.7% |  | 18.9% | 15.9% |  | 38.1% | 8.5% |  | 11.8% | 12.5% |  |
| S1Q3T3 | 19% | 17.3% | 20.8% | 0.4 | 24.3% | 15.9% | 0.2 | 23.8% | 25.5% | 0.6 | 29.4% | 25.0% | 0.5 |
| Premature ventricular complexes | 23% | 15.4% | 31.3% | 0.050 | 21.6% | 23.8% | 0.5 | 14.3% | 23.4% | 0.3 | 29.4% | 8.3% | 0.1 |
| Premature atrial complexes | 13% | 5.8% | 20.8% | 0.02 | 8.1% | 15.9% | 0.2 | 4.8% | 12.8% | 0.3 | 23.5% | 4.2% | 0.1 |
| Poor R wave progression | 14% | 17.3% | 10.4% | 0.2 | 18.9% | 11.1% | 0.2 | 14.3% | 14.9% | 0.6 | 5.9% | 16.7% | 0.3 |
| Rhythm on admission |  |  |  |  |  |  |  |  |  |  |  |  |  |
| Sinus rhythm | 56% | 57.7% | 54.2% | 0.9 | 70.3% | 47.6% | 0.059 | 66.7% | 46.8% | 0.3 | 52.9% | 58.3% | 0.9 |
| Sinus tachycardia | 33% | 30.8% | 35.4% |  | 18.9% | 41.3% |  | 28.6% | 42.6% |  | 35.3% | 29.2% |  |
| Atrial fibrillation | 11% | 11.5% | 10.4% |  | 10.8% | 11.1% |  | 4.8% | 10.6% |  | 11.8% | 12.5% |  |
| Rhythm during admission |  |  |  |  |  |  |  |  |  |  |  |  |  |
| Atrial arrhythmias | 29% | 17.3% | 27.1% | 0.2 | 16.2% | 25.4% | 0.2 | 19.0% | 25.5% | 0.4 | 23.5% | 25.0% | 0.6 |
| Atrial fibrillation | 18% | 15.4% | 20.8% | 0.3 | 13.5% | 20.6% | 0.3 | 14.3% | 23.4% | 0.3 | 23.5% | 20.8% | 0.6 |
| Atrial flutter | 6% | 5.8% | 6.3% | 0.6 | 5.4% | 6.3% | 0.6 | 4.8% | 2.1% | 0.5 | 5.9% | 8.3% | 0.6 |
| Atrial tachycardia | 5% | 1.9% | 8.3% | 0.5 | 2.7% | 3.2% | 0.7 | 4.8% | 4.3% | 0.7 | 0.0% | 4.2% | 0.6 |
| Sinus bradycardia | 13% | 17.3% | 8.3% | 0.1 | 24.3% | 6.3% | 0.01 | 9.5% | 10.6% | 0.6 | 5.9% | 16.7% | 0.3 |
| Sinus tachycardia | 81% | 78.8% | 83.3% | 0.4 | 67.6% | 88.9% | 0.01 | 90.5% | 80.9% | 0.3 | 82.4% | 75.0% | 0.4 |
| Left atrial abnormality | 5.1% | 9.8% | 0% | 0.03 | 5.6% | 4.8% | 0.6 | 0.0% | 4.3% | 0.5 | 12.5% | 4.2% | 0.3 |
| Right atrial abnormality | 7.1% | 5.9% | 8.5% | 0.4 | 5.6% | 8.1% | 0.5 | 5.0% | 8.7% | 0.5 | 6.3% | 12.5% | 0.5 |
| Left axis deviation | 24.2% | 21.6% | 27.1% | 0.8 | 27.0% | 22.6% | 0.8 | 19.0% | 27.7% | 0.4 | 17.6% | 33.3% | 0.2 |
| Right axis deviation | 2.1% | 2% | 2.1% |  | 2.7% | 1.6% |  | 0.0% | 4.3% |  | 0.0% | 0.0% |  |
| Right bundle branch block (RBBB) | 4% | 3.8% | 4.2% | 0.7 | 5.4% | 3.2% | 0.5 | 4.8% | 4.3% | 0.7 | 5.9% | 0.0% | 0.4 |
| Incomplete RBBB | 3% | 3.8% | 2.1% | 0.5 | 0.0% | 4.8% | 0.2 | 0.0% | 2.1% | 0.7 | 0.0% | 4.2% | 0.6 |
| Q waves | 27% | 28.8% | 25% | 0.4 | 35.1% | 22.2% | 0.1 | 23.8% | 34.0% | 0.3 | 35.3% | 37.5% | 0.6 |
| Inferior Q waves | 22% | 21.2% | 22.9% | 0.5 | 21.6% | 22.2% | 0.02 | 23.8% | 25.5% | 0.6 | 29.4% | 29.2% | 0.8 |
| Anterior Q waves | 2% | 3.8% | 0% |  | 5.4% | 0.0% |  | 0.0% | 2.1% |  | 5.9% | 4.2% |  |
| Q waves in ≥ 2 territories | 3% | 3.8% | 2.1% |  | 8.1% | 0.0% |  | 0.0% | 6.4% |  | 0.0% | 4.2% |  |
| Low voltage | 16% | 9.6% | 22.9% | 0.06 | 13.5% | 17.5% | 0.4 | 14.3% | 14.9% | 0.6 | 5.9% | 29.2% | 0.06 |
| Left ventricular hypertrophy | 15% | 17.3% | 12.5% | 0.3 | 16.2% | 14.3% | 0.5 | 9.5% | 19.1% | 0.3 | 17.6% | 12.5% | 0.5 |
| Left anterior fascicular block | 12% | 13.5% | 10.4% | 0.4 | 18.9% | 7.9% | 0.09 | 9.5% | 17.0% | 0.3 | 5.9% | 20.8% | 0.2 |
| Bi-fascicular block | 4% | 3.8% | 4.2% | 0.7 | 5.4% | 3.2% | 0.5 | 4.8% | 4.3% | 0.7 | 5.9% | 0.0% | 0.4 |
| Normal QRS width | 95% | 94.2% | 95.8% | 0.5 | 94.6% | 95.2% | 0.6 | 95.2% | 95.7% | 0.67 | 94.1% | 100% | 0.4 |

**Table 3S:** Correlation of EKG findings with median D-dimer of 1 mg/L, median C-reactive protein (CRP) of 175 mg/l, median Ferritin level of 900 ng/ml and median Length of stay (LOS) of 10 days

| EKG finding | D-Dimer |  | P-value | CRP |  | P-value | Ferritin |  | P-value | LOS |  | P-value |
| --- | --- | --- | --- | --- | --- | --- | --- | --- | --- | --- | --- | --- |
|  | <1 | ≥1 |  | <175 | ≥175 |  | <900 | ≥900 |  | <10 | ≥10 |  |
| 1<sup>st</sup> degree heart block (HB) | 6.3% | 11.5% | 0.47 | 13.6% | 11.4% | 0.5 | 13.6% | 12.2% | 0.5 | 10.6% | 13.2% | 0.46 |
| Transient 1<sup>st</sup> degree HB | 0.0% | 7.7% | 0.4 | 9.1% | 2.3% | 0.2 | 9.1% | 4.1% | 0.5 | 6.4% | 5.7% | 0.7 |
| Persistent 1<sup>st</sup> degree HB | 6.3% | 3.8% |  | 4.5% | 9.1% |  | 4.5% | 8.2% |  | 4.3% | 7.5% |  |
| Transient ST elevation | 0.0% | 7.7% | 0.7 | 2.3% | 6.8% | 0.7 | 2.3% | 8.2% | 0.5 | 4.3% | 5.7% | 0.7 |
| Persistent ST elevation | 6.3% | 3.8% |  | 2.3% | 2.3% |  | 2.3% | 4.1% |  | 2.1% | 3.8% |  |
| ST depression | 6.3% | 9.6% |  | 9.1% | 9.1% |  | 9.1% | 8.2% |  | 6.4% | 9.4% |  |
| Early repolarization | 0.0% | 1.9% |  | 2.3% | 0.0% |  | 2.3% | 0.0% |  | 2.1% | 0.0% |  |
| T wave inversion | 37.5% | 48.1% | 0.3 | 36.4% | 52.3% | 0.09 | 36.4% | 51.0% | 0.1 | 46.8% | 37.7% | 0.2 |
| T wave inversion (Inferior) | 0.0% | 5.8% | 0.7 | 2.3% | 9.1% | 0.2 | 2.3% | 8.2% | 0.3 | 4.3% | 5.7% | 0.1 |
| (Anterior) | 18.8% | 19.2% |  | 15.9% | 20.5% |  | 15.9% | 20.4% |  | 12.8% | 22.6% |  |
| (Lateral) | 0.0% | 3.8% |  | 0.0% | 4.5% |  | 0.0% | 4.1% |  | 2.1% | 1.9% |  |
| (≥ two territories) | 18.8% | 19.2% |  | 18.2% | 18.2% |  | 18.2% | 18.4% |  | 27.7% | 7.5% |  |
| S1Q3T3 | 12.5% | 19.2% | 0.4 | 18.2% | 22.7% | 0.3 | 18.2% | 22.4% | 0.4 | 21.3% | 17.0% | 0.4 |
| Premature ventricular complexes | 12.5% | 26.9% | 0.2 | 25.0% | 18.2% | 0.3 | 25.0% | 18.4% | 0.3 | 25.5% | 20.6% | 0.4 |
| Premature atrial complexes | 6.3% | 15.4% | 0.3 | 15.9% | 9.1% | 0.3 | 15.9% | 10.2% | 0.3 | 12.8% | 13.2% | 0.9 |
| Poor R wave progression | 18.8% | 11.5% | 0.3 | 18.2% | 11.4% | 0.2 | 18.2% | 10.2% | 0.2 | 17.0% | 11.3% | 0.2 |
| Rhythm on admission |  |  |  |  |  |  |  |  |  |  |  |  |
| Sinus rhythm | 56.3% | 59.6% | 0.1 | 61.4% | 45.5% | 0.2 | 61.4% | 46.9% | 0.34 | 59.6% | 52.8% | 0.7 |
| Sinus tachycardia | 25.0% | 36.5% |  | 27.3% | 43.2% |  | 27.3% | 40.8% |  | 29.8% | 35.8% |  |
| Atrial fibrillation | 18.8% | 3.8% |  | 11.4% | 11.4% |  | 11.4% | 12.2% |  | 10.6% | 11.3% |  |
| Rhythm during admission |  |  |  |  |  |  |  |  |  |  |  |  |
| Atrial arrhythmia | 18.8% | 17.3% | 0.57 | 20.5% | 20.5% | 0.6 | 20.5% | 22.4% | 0.5 | 14.9% | 28.3% | 0.08 |
| Atrial fibrillation | 18.8% | 13.5% | 0.4 | 20.5% | 18.2% | 0.5 | 20.5% | 16.3% | 0.4 | 12.8% | 22.6% | 0.1 |
| Atrial flutter | 6.3% | 1.9% | 0.4 | 2.3% | 6.8% | 0.3 | 2.3% | 8.2% | 0.2 | 4.3% | 7.5% | 0.3 |
| Atrial tachycardia | 6.3% | 3.8% | 0.5 | 0.0% | 4.5% | 0.2 | 0.0% | 6.1% | 0.1 | 0.0% | 5.7% | 0.1 |
| Sinus bradycardia | 18.6% | 9.6% | 0.2 | 18.2% | 9.1% | 0.1 | 18.2% | 8.2% | 0.1 | 12.8% | 13.2% | 0.5 |
| Sinus tachycardia | 81.3% | 92.3% | 0.2 | 72.7% | 90.9% | 0.02 | 72.7% | 89.8% | 0.03 | 74.5% | 86.8% | 0.1 |
| Left atrial abnormality | 6.3% | 2.0% | 0.4 | 2.4% | 9.1% | 0.1 | 2.4% | 8.2% | 0.2 | 2.2% | 7.5% | 0.2 |
| Right atrial abnormality | 0.0% | 7.8% | 0.3 | 11.9% | 4.5% | 0.1 | 11.9% | 4.1% | 0.1 | 6.7% | 7.5% | 0.5 |
| Left axis deviation | 31.3% | 23.1% | 0.7 | 23.3% | 31.8% | 0.4 | 23.3% | 28.6% | 0.8 | 25.5% | 23.1% | 0.9 |
| Right axis deviation | 0.0% | 1.9% |  | 2.3% | 0.0% |  | 2.3% | 2.0% |  | 2.1% | 1.9% |  |
| Right bundle branch block (RBBB) | 6.3% | 1.9% | 0.4 | 6.8% | 2.3% | 0.3 | 6.8% | 2.0% | 0.3 | 4.3% | 3.8% | 0.6 |
| Incomplete RBBB | 6.3% | 1.9% | 0.4 | 2.3% | 4.5% | 0.5 | 2.3% | 4.1% | 0.5 | 2.1% | 3.8% | 0.5 |
| Q wave | 18.8% | 23.1% | 0.5 | 27.3% | 29.5% | 0.5 | 27.3% | 28.6% | 0.5 | 25.5% | 28.3% | 0.4 |
| Inferior Q wave | 12.5% | 23.1% | 0.1 | 18.2% | 29.5% | 0.1 | 18.2% | 28.6% | 0.1 | 19.1% | 24.5% | 0.1 |
| Anterior Q wave | 0.0% | 0.0% |  | 2.3% | 0.0% |  | 2.3% | 0.0% |  | 0.0% | 3.8% |  |
| Q wave in ≥ 2 territories | 6.3% | 0.0% |  | 6.8% | 0.0% |  | 6.8% | 0.05 |  | 6.4% | 0.0% |  |
| Low voltage | 12.5% | 15.4% | 0.56 | 18.2% | 11.4% | 0.3 | 18.2% | 12.2% | 0.3 | 23.4% | 9.4% | 0.051 |
| Left ventricular hypertrophy | 6.3% | 11.5% | 0.4 | 6.8% | 25.0% | 0.01 | 6.8% | 22.4% | 0.03 | 19.1% | 11.3% | 0.2 |
| Left anterior fascicular block | 25.0% | 3.8% | 0.02 | 13.6% | 13.6% | 0.6 | 13.6% | 12.2% | 0.5 | 17.0% | 7.5% | 0.1 |
| Left posterior fascicular block | 0.0% | 1.9% | 0.7 | 0.0% | 0.0% | - | 0.0% | 2.0% | 0.5 | 0.0% | 1.9% | 0.5 |
| Bi-fascicular block | 6.3% | 1.9% | 0.4 | 6.8% | 2.3% | 0.3 | 6.8% | 2.0% | 0.2 | 4.3% | 3.8% | 0.6 |
| Normal QRS width | 93.8% | 98.1% | 0.4 | 90.9% | 97.7% | 0.1 | 90.9% | 98.0% | 0.1 | 95.7% | 94.3% | 0.5 |

For our analysis, categorical and continuous variables were reported as frequency or percentage and mean ± standard error (SE) or median ± interquartile range, based on the normality of the data. Baseline demographics and comorbidities between groups were compared using the Pearson χ^2^ test for categorical variables and one-way linear regression for continuous variables (Table 4S supplementary index)

**Table 4S:** Baseline characteristics of our cohort of 100 patients diagnosed with COVID-19

| Variable | N= 100 | Survivors<br>N=52 | Non-survivors<br>N=48 | P-value |
| --- | --- | --- | --- | --- |
| Age (median, IQR) | 66 (57-74) | 65 (47.5-71) | 68.5 (60-75) | 0.1 |
| Race |  |  |  |  |
| Male gender | 61% | 67.3% | 54.2% | 0.2 |
| African Americans | 77% | 82.7% | 70.8% | 0.3 |
| White | 6% | 3.8% | 8.3% |  |
| Other | 17% | 13.5% | 20.8% |  |
| Body mass index kg/m<sup>2</sup> | 31 (26.1 -39 ) | 30.9 (25.9-36.8) | 31.2 (26.6-41) | 0.2 |
| Hypertension | 71% | 71.2% | 70.8% | 1.0 |
| Diabetes Mellitus | 40% | 30.8% | 50% | 0.05 |
| Dyslipidemia | 41% | 40.4% | 41.7% | 0.9 |
| Chronic/end-stage renal disease | 19% | 21.2% | 16.7% | 0.6 |
| Lung disease | 26% | 23.1% | 29.2% | 0.5 |
| Obstructive sleep apnea | 11% | 5.8% | 16.7% | 0.08 |
| Cancer | 15% | 13.5% | 16.7% | 0.6 |
| Immunological | 7% | 7.6% | 6.2% | 0.6 |
| Mental disorder | 12% | 15.3% | 8.3% | 0.3 |
| Neurological disorder | 11% | 11.5% | 10.4% | 0.9 |
| Coronary artery disease | 12% | 17.3% | 6.3% | 0.08 |
| Systolic heart failure | 10% | 15.4% | 4.2% | 0.06 |
| Atrial fibrillation/Atrial flutter | 10% | 15.4% | 4.2% | 0.06 |
| Troponin (Peak) | 38.5(10-187.2) | 23.5 ( 9- 185) | 43.5 (18.75 -187.2) | 0.2 |
| Brain Natriuretic peptide (BNP) | 130 (52.5-411.5) | 123 ( 35.25 -261 ) | 158 ( 56 -1034 ) | 0.5 |
| Ferritin | 971 ( 418-1952.5) | 884 ( 383-1345) | 1035 (522-2802) | 0.04 |
| D-dimer | 2.2 ( 1.12 -21.35) | 1.34 ( 0.5-4.3) | 4.9 ( 1.6 -35.2 ) | 0.002 |
| C-reactive protein (CRP) | 175.0 (81.7-292.0) | 171.5 (66.5-267.5) | 207.5 (95.7-323.0) | 0.2 |
| Treatment |  |  |  |  |
| No Hydroxychloroquine (HY)/<br>Azithromycin(AZ)/Doxycycline | 12 (12.0%) | 4 (7.7%) | 8 (16.7%) | 0.02 |
| HY only | 19 (19.0%) | 9 (17.3%) | 10 (20.8%) |  |
| HY & AZ | 40 (40.0%) | 17 (32.7%) | 23 (47.9%) |  |
| HY & Doxycycline | 29 (29.0%) | 22 (42.3%) | 7 (14.6%) |  |
| Tocilizumab | 5% | 0% | 10.4% | 0.02 |
| Steroids | 53% | 50% | 56.3% | 0.5 |
| L-ascorbic acid | 24% | 23.1% | 25% | 0.8 |
| Zinc | 16% | 15.4% | 16.7% | 0.9 |
| Anticoagulation | 32% | 25% | 39.6% | 0.1 |
| Single antiplatelet drug | 33% | 44.2% | 20.8% | 0.03 |
| Dual antiplatelet therapy | 7% | 3.8% | 10.4% |  |
| Statins | 38% | 38.5% | 37.5% | 0.9 |
| ≥ Vasopressor | 37% | 5.8% | 70.8% | <0.001 |
| Beta blockers | 41% | 50% | 31.3% | 0.06 |
| Calcium channel blocker | 8% | 7.7% | 8.3% | 0.6 |
| Amiodarone | 7% | 3.8% | 10.4% | 0.2 |
| Corrected QT(QTc) on admission (milliseconds) | 443 ( 410.2 -472.7) | 435 ( 404.5-476.7 ) | 447 (416.2 -465.7) | 0.8 |
| Smoking | 30% | 36.5% | 22.9% | 0.1 |
| Alcohol | 20% | 23.1% | 16.7% | 0.4 |
| Illicit drugs | 9% | 15.4% | 2.1% | 0.02 |
| Potassium trough | 3.6 (3.3-4.0 ) | 3.7 ( 3.3-3.9) | 3.6 (3.4-4 ) | 0.4 |
| Potassium peak | 4.75 (4.32-5.5) | 4.75 (4.3-5.5) | 4.75 ( 4.4-5.4) | 0.7 |
| Magnesium trough | 1.9 (1.7-2.1) | 1.9 ( 1.72-2.1) | 2 (1.7-2.2) | 0.5 |
| Magnesium peak | 2.4 (2.2-2.7) | 2.3 (2.1-2.6) | 2.6 (2.3- 2.87) | 0.001 |
| Sodium trough | 134 (131-137) | 134(132-136) | 135 ( 130-138) | 0.5 |
| Sodium peak | 143(140-147) | 142 ( 140-145) | 145 ( 140.2 -148.7) | 0.03 |
| Calcium trough | 7.9(7.4-8.3) | 8.1(7.6-8.4) | 7.65 (7.1-8.2) | 0.005 |
| Calcium peak | 9.0(8.5 -9.4) | 9.2 ( 8.9-9.6) | 8.7 ( 8.5-9.2) | 0.001 |
| Phosphorus trough | 2.7 (2.2 -3.8) | 2.6 (2.1-3.2) | 3 ( 2.2-4.3) | 0.2 |
| Phosphorus peak | 4.5( 3.4-6.7) | 3.7( 3-4.7) | 5.3(4.1-7.7) | <0.001 |
| Intensive care unit (ICU) admission | 48 (48.0%) | 8 (21.6%) | 40 (63.5%) | <0.001 |
| ICU length of stay | 3 ( 0- 8.9) | 0 (0 – 3.75) | 7 ( 2.2 -13.1) | < 0.001 |
| Hospital length of stay | 10.4 ( 6.7-16) | 10.6 (7.07 -16.4) | 9.4 ( 5.7-15.6) | 0.5 |

Our cohort included 61% males and 77% African Americans. The median age and BMI were 66 years (57- 74) and 31 kg/m^2^ (26.1-39), respectively. History of Hypertension, Diabetes Mellitus, Cancer and lung disease was present in 71%, 40%, 15% and 26% of the patients, respectively. 12% had coronary artery disease, 10% had Systolic heart failure and 12% had history of atrial arrhythmias at baseline. The median hospital stay was 10.4 days (6.7-16) with 48% mortality.

We observed atrial arrhythmias in 29% of the patients (17% new onset), First degree heart block in 12% (6% were transient), ST-T segment changes in 17%, S1Q3T3 pattern in 19%, Premature ventricular complexes (PVCs) in 23%, premature atrial complexes(PACs) in 13%, Q waves in 27%, T wave inversion in 42% of the cases. Long QTc (> 470 in males, >480 in females) was noted in 37 cases, of whom 20 patients were on Hydroxychloroquine and Azithromycin. There was no correlation between long QTc and mortality (P = 0.5).

Myocardial injury due to hypoxia and direct viral tissue invasion, sympathetic hyperactivity secondary to the cytokine storm and drug induced QTc prolongation are postulated mechanism of arrhythmias in COVID-19 cases (2)

Transient complete heart block was previously reported (3, 4).In our cohort, only first degree heart block was present without higher grade block.

S1Q3T3 pattern, possibly due to cor pulmonale, was reported in 2 Covid-19 cases (4). It was noted in 19% of patients in our cohort and did not correlate with mortality (P = 0.4)

ST elevation without obstructive coronary artery disease was reported in COVID-19 patients (5). We noted 8 cases of ST elevation, of which 5 were transient and 8 cases of ST depression. None of these patients underwent coronary angiogram during hospitalization. ST segment changes did not correlate with mortality (P = 0.5)

Premature complexes are possibly explained by the higher sympathetic tone, vasopressors use, electrolyte abnormalities and myocarditis. Right ventricular outflow tract was the most common site of origin (47% of cases). While PACs correlated with mortality (P = 0.02), PVCs correlation didn’t reach statistical significance (P = 0.05). EKG abnormalities are prevalent in this cohort of COVID-19 patients and can be a helpful diagnostic tool for underlying cardiac pathologies.

## Data Availability

All relevant data are included in the manuscript

## Acknowledgements

None.

## Financial Disclosures

Authors have no financial disclosures to report.

## Conflict of Interest

The authors declare that they have no conflict of interest concerning this article.

## Informed Consent

Not applicable.

## Authors Contributions

Shokr M designed the research, collected the data and wrote the manuscript. Chehab O analyzed the data. All authors contributed to data collection. Shokr M and Lieberman R approved the final version of the manuscript.

## Notes

### Competing Interest Statement

The authors have declared no competing interest.

### Funding Statement

No external funding was received.

### Author Declarations

Approved by the IRB committee at Wayne State University, Detroit, MI, USA.

